# Detection of Anti-Non-α-Gal Xenoreactive Antibodies in Human Blood Products

**DOI:** 10.1101/2024.11.26.24318030

**Authors:** Joseph M. Ladowski, Meghan Hu, Janghoon Yoon, Zheng Chen, Stuart Knechtle, Annette Jackson, Jean Kwun

**Affiliations:** Duke Transplant Center, Department of Surgery, Duke University School of Medicine, Durham, NC; Department of Surgery, Duke University School of Medicine, Durham, NC; Clinical Transplantation Immunology Laboratory, Duke University School of Medicine, Durham, NC

**Author notes:** **To whom correspondence should be addressed:** Joseph Ladowski MD PhD, 2301 Erwin Rd, DUMC Box 3704, Durham, North Carolina 27710., Jean Kwun, PhD, 210 Research Drive, GSRBII, Room 4006C, DUMC Box 2645, Durham, NC 27710, USA. Both authors contributed equally as co-first authors (Joseph M. Ladowski, Meghan Hu). Reflects shared senior authorship (Annette Jackson, Jean Kwun).

**Keywords:** Xenoreactive antibody, Fresh frozen plasma, Cryoprecipitate, Xenotransplantation, Alloantibody, Kidney transplantation

## Abstract

**Background:** Surgical bleeding is a risk in any solid organ transplant, and commonly addressed with the transfusion of human blood product to replace or supplement coagulation factors. It is unknown if these blood products would harm xenotransplanted pig organs in human recipients demonstrating coagulopathy. The aim of this study was to investigate *in vitro* if blood products such as fresh frozen plasma (FFP) or cryoprecipitate (cryo) contain xenoantibodies capable of cytotoxicity to GTKO pig cells.

**Methods:** We obtained 12 individual single donor (7 FFP and 5 cryo) blood products from our institution’s blood bank for testing. PBMCs were obtained from a GTKO/hCD55 pig for use as target cells. We performed a series of flow cytometry crossmatch and complement-dependent cytotoxicity assays.

**Results:** We found that all the tested blood products contained some degree of IgM and IgG xenoantibody. Tests using a 1:50 dilution revealed a significant decrease in IgM xenoantibody binding, but an increase the detection of IgG binding. Multiple preparations were capable of GTKO PBMC cytotoxicity but the level of antibody binding and cell death varied by preparation.

**Conclusions:** Both FFP and cryo contain IgM and IgG non-αGal xenoantibodies capable of killing GTKO PBMCs, though the level varies by preparation. While some centers utilize a genetic background with mutations in the three enzymes responsible for the known xenoantigens, others are investigating the GTKO pig as a potential option. These results suggest that a center pursuing a human xenotransplantation study with a GTKO genetic background should pre-screen blood products prior to administration.

## Introduction

Xenotransplantation of genetically engineered pig organs to human recipients,has long been a theoretical option to increase the donor organ pool(1).The initial barrier to human clinical xenotransplantation, specifically hyperacute rejection, appears to have been overcome by multiple groups by combining genetically engineered donor pigs with enhanced immunosuppression(2–6). As the field considers a clinical trial, focus is now being placed on the perioperative care of xenograft recipients – including the potential passive transfusion of detrimental xenoantibodies from blood products such as intravenous immunoglobulin (IVIg) (7– 9). While the prevalence of xenoantibodies in IVIg may be debated, it represents one of many options to supplement the immune system of transplant recipients. However, the available options to treat coagulopathy and bleeding in transplant recipients are more limited, with increased reliance on the use of human blood products. This is particularly relevant to cardiac transplant recipients, of whom up to 90% may require a blood product in the first week post-transplant(10,11).

The coagulation system is a complex balance between clotting and fibrinolysis and consists of fibrinogen, von Willebrand factor, and multiple clotting factors(12). Hemorrhage, such as that experienced in a major surgical procedure like a solid organ transplant, can disrupt this balance. Coagulopathies resulting from surgical hemorrhage are often addressed with replacement of clotting factors via either fresh frozen plasma (FFP) or cryoprecipitate (cryo) derived from healthy human donors to provide supplemental fibrinogen, clotting factors, and von Willebrand factor(12,13). While FFP units are derived from a single donor, cryo can potentially be prepared from 4-6 pooled donors (14). FFP has been shown to contain antibodies that may result in cellular damage. For example, ABO compatibility is a requirement for FFP administration given the known risk of hemolytic transfusion reaction secondary to transfusion of anti-ABO antibodies (15,16). While it has been established that the majority of individuals possess non-galactose-α1,3-galactose (αGal) xenoantibodies, it is unknown if blood products derived from a healthy individual possess xenoantibodies capable of recognizing GTKO cells(17).

We sought to examine the presence and prevalence of non-αGal xenoantibodies in the blood products commonly administered post-operatively to treat surgical coagulopathy: FFP and cryo. Additionally, we investigated the potential pathogenicity of these xenoantibodies in a complement-dependent cytotoxicity (CDC) assay. Our results confirm the presence of cytotoxic non -αGal xenoantibodies in blood products that may be a detriment if passively transferred into a xenograft recipient.

## Materials and Methods

### Institutional Approval

Any animal care was conducted in accordance with National Institutes of Health (NIH) guidelines with approval by the Duke University Institutional Animal Care and Use Committee (Duke IACUC A032-20-02 and 093-23-04). The use of discarded blood products from the Duke Blood Bank was determined to be exempt by the Duke University Institutional Review Board (IRB Pro00115735).

### Sample Collection

Cryopreserved peripheral blood mononuclear cells (PBMCs) stored at -80C in 90% fetal bovine serum and 10% dimethyl sulfoxide (Thermofisher Scientific, Waltham, MA) from a GGTA1/CD55Tg (1KO/1TG) background pigs (National Swine Resource and Research Center (NSRRC; University of Missouri-Columbia, Columbia, MO)) were used in this study. MHC typing was performed as described previously by the Gift of Hope Organ & Tissue Donor Network with high-resolution SLA typing imputed based on previously published SLA nomenclature reports (18–19).

Discarded fresh frozen plasma (FFP) and cryoprecipitate (cryo) samples were obtained from the Duke University Blood Bank. The FFP were from a single donor and the cryo samples were pooled from five donors, as per hospital protocols. In the case of FFP, the samples were aliquoted and frozen at -80°C until use. The cryo samples were stored at room temperature (due to significant precipitate formation when frozen) and used within 3 months of expiration date. Internal testing showed no significant degradation of DSA during this three-month time period (data not shown). At the time of testing, all sera were thawed and heat-inactivated at 57°C for 30 minutes to degrade residual complement that may confound the analysis.

### Flow Cytometry Crossmatch

To examine for presence of XAbs, the FCXM protocol was performed as previously described with minor modifications (20,21). Briefly, 2.0 ×10^5^ PBMCs from the donor pig were thawed and stained with Live/Dead Fixable Blue (Thermofisher Scientific, Waltham, MA) at the manufacturer recommended concentration for 30 minutes at room temperature. The cells were then washed, plated in a 96-well round bottom culture plate (Corning, New York, New York City, USA), and blocked with Goat IgG whole molecule solution (Jackson Immuno Research Laboratory, Inc.:005-000-003) for 15 minutes at 4ºC. The cells were washed and resuspended in 50 uL of the respective blood product at either a “neat” concentration or diluted 1:50 with 1X PBS. The cells were incubated for 30 minutes at 4ºC, a length of time and temperature previously chosen to provide internally consistent results and theoretically maximize IgM binding. The PBMCs were selected based on sample availability and maximal viability.

Following antibody binding, the cells were washed and stained with anti-human IgM FITC (Clone G20-127, BD Pharmingen, KPL, Gaithersburg, MD), anti-human IgG BV421 (Clone G18-145, BD Biosciences), anti-swine CD3e mAb (BB23-8E6-8C8, BD, San Jose, CA), and anti-human CD21 mAb (B-ly4, BD, San Jose, CA) at recommended dilutions. A negative control of cells stained for CD21, CD3, IgM and IgG without blood product was used to determine the background antibody binding and autofluorescence. To control for target cell size as a variable, values are reported as MFI relative to negative control. Flow cytometric analysis was performed on a BD LSRFortessa. Gating strategy for the flow cytometry crossmatch was based on size, singlet population, viability as determined by low uptake of Live/Dead Fixable Blue, lymphocyte gate as determined by FSC vs SSC, and in some cases CD3+ or CD21+ expression. The median fluorescence intensity (MFI) of IgM or IgG was measured and all analysis was performed using FlowJo software version 10.0 (Tree Star, Ashland, OR). MFI values are reported either as average MFI for samples assessed ± the standard deviation or MFI fold over background.

### Complement-Dependent Cytotoxicity Assay

A complement-dependent cytotoxicity (CDC) assay was performed as previously described with minor modifications (20). Briefly, 2.0 ×10^5^ PBMCs from the donor pig were thawed and resuspended in Hank’s Balanced Salt Solution (HBSS, Thermofisher Scientific, Waltham, MA), and added to individual wells in a 96-well round bottom culture plate. The cells were pelleted by centrifugation at 500 x g for 5 minutes at room temperature, decanted, and incubated with 100 uL of the respective blood product at either a “neat” concentration or a 1:50 dilution. Half of the samples were incubated with respective blood product that underwent diothiothreitol (DTT, Thermofisher Scientific, Waltham, MA) treatment with a 2.5 mM concentration of DTT for 30 minutes at 37°C to disrupt IgM antibodies.

The cells were washed three times with 1X HBSS and treated with 50 uL/well of an 11-fold dilution of Low-Tox H Rabbit Complement (Cedarlane, Burlington, NC) for 90 minutes at 37°C. The cells were washed an additional two times with 1X HBSS and stained with Live/Dead Fixable Blue stain per the manufacturer’s recommended concentration for 30 minutes at room temperature. Two additional washes were performed, and the cells were analyzed by flow cytometry. Three negative controls were performed: cells incubated with media alone, cells incubated with rabbit complement alone, and cells incubated with sera alone were used to determine the background level of cytotoxicity. A positive control of cells with 70% ethanol was used to confirm appropriate cellular death. CDC samples were analyzed on the same BD LSRFortessa and gated based on size, singlet population, and viability as determined by Live/Dead Fixable Blue. The results are depicted as cellular cytotoxicity or killing over background levels.

### Statistical Analysis

All statistical analyses were performed using Prism 9.0 (GraphPad Software, San Diego, CA). Mann-Whitney non-parametric unpaired test was used to compare antibody binding between sensitized and non-sensitized recipient sera samples. Values of p < 0.05 were considered to be significant.

## Results

To determine if human blood products contained xenoantibodies capable of binding pig cells, we tested seven (7) FFP and five (5) cryo individual samples with a flow cytometry crossmatch (FCXM) with PBMCs from GTKO pigs. The samples demonstrated varying levels of xenoantibody binding with statistically more IgM than IgG antibody binding to both CD3^+^ T cells (relative IgM MFI 44.43 ± 38.83 vs relative IgG MFI 3.003 ± 2.567, p = 0.0002) and CD21^+^ B cells (relative IgM MFI 24.37 ± 9.159 vs relative IgG MFI 2.357 ± 2.206, p = 0.0003) (**Figure 1)** in PBMCs. A 1:50 dilution was performed prior to FCXM to investigate a potential prozone effect or blocking effect by a xenoantibody of lower concentration but higher avidity for the xenoantigen. This resulted in a significant decrease in IgM xenoantibody binding to CD3+ (relative IgM MFI 44.43 ± 38.83 vs 4.165 ± 0.9187, p = 0.0017) and CD21+ PBMCs (mean MFI 24.37 ± 9.159 vs 3.453 ± 1.021, p < 0.0001). There was no statistically significant change in IgG xenoantibody binding to CD3+ (mean MFI 3.003 ± 2.567 vs 2.144 ± 1.083, p >0.9999) but the 1:50 dilution resulted in a statistically significant increase in IgG binding to CD21+ PBMCs (mean MFI 2.357 ± 2.206 vs 2.940 ± 1.084, p = 0.0266). Individual blood product sample variability in xenoantibody binding can be found in **Figure 2**.

**Figure 1:**
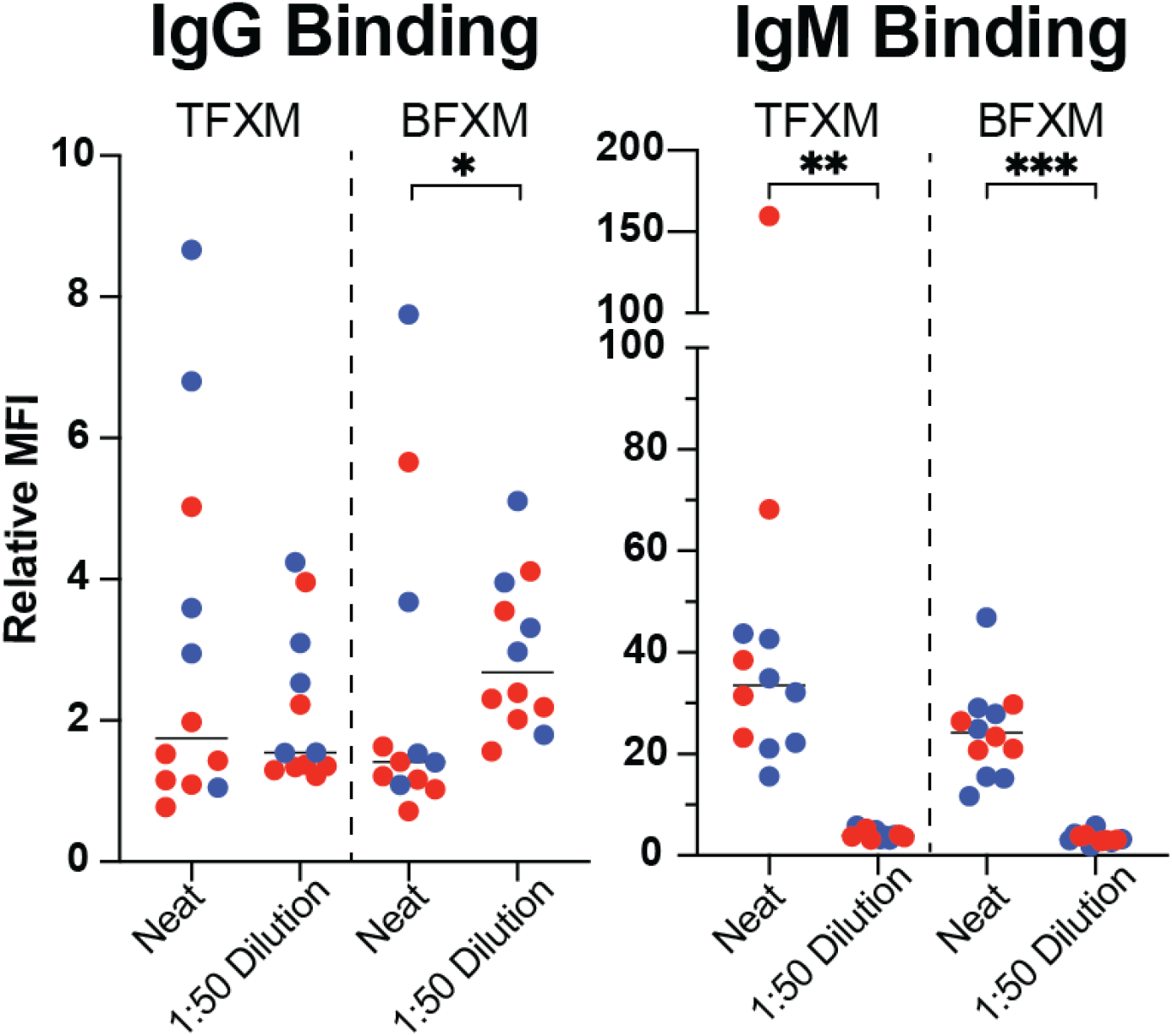
Detection of xenoantibodies in blood products. Shown are the relative IgM and IgG xenoantibody binding of seven FFP (red circles) and five cryo (blue circles) samples to GTKO PBMCs. Statistical significance was noted for IgM xenoantibody binding to CD3+ vs CD21+ lymphocytes (left figure, p = 0.0269) but not IgG xenoantibody binding to CD3+ vs CD21+ lymphocytes (right figure, p = 0.1514). A dilutional effect was explored with a 1:50 dilution which resulted in a significant decrease of IgM xenoantibody binding to both CD3+ (p = 0.0017) and CD21+ (p = 0.0005) PBMCs, but a non-significant for IgG antibody binding to CD3+ (p > 0.9999) and significant increase in binding to CD21+ PBMCs (p = 0.0266). All values are depicted as MFI relative to negative control.

**Figure 2:**
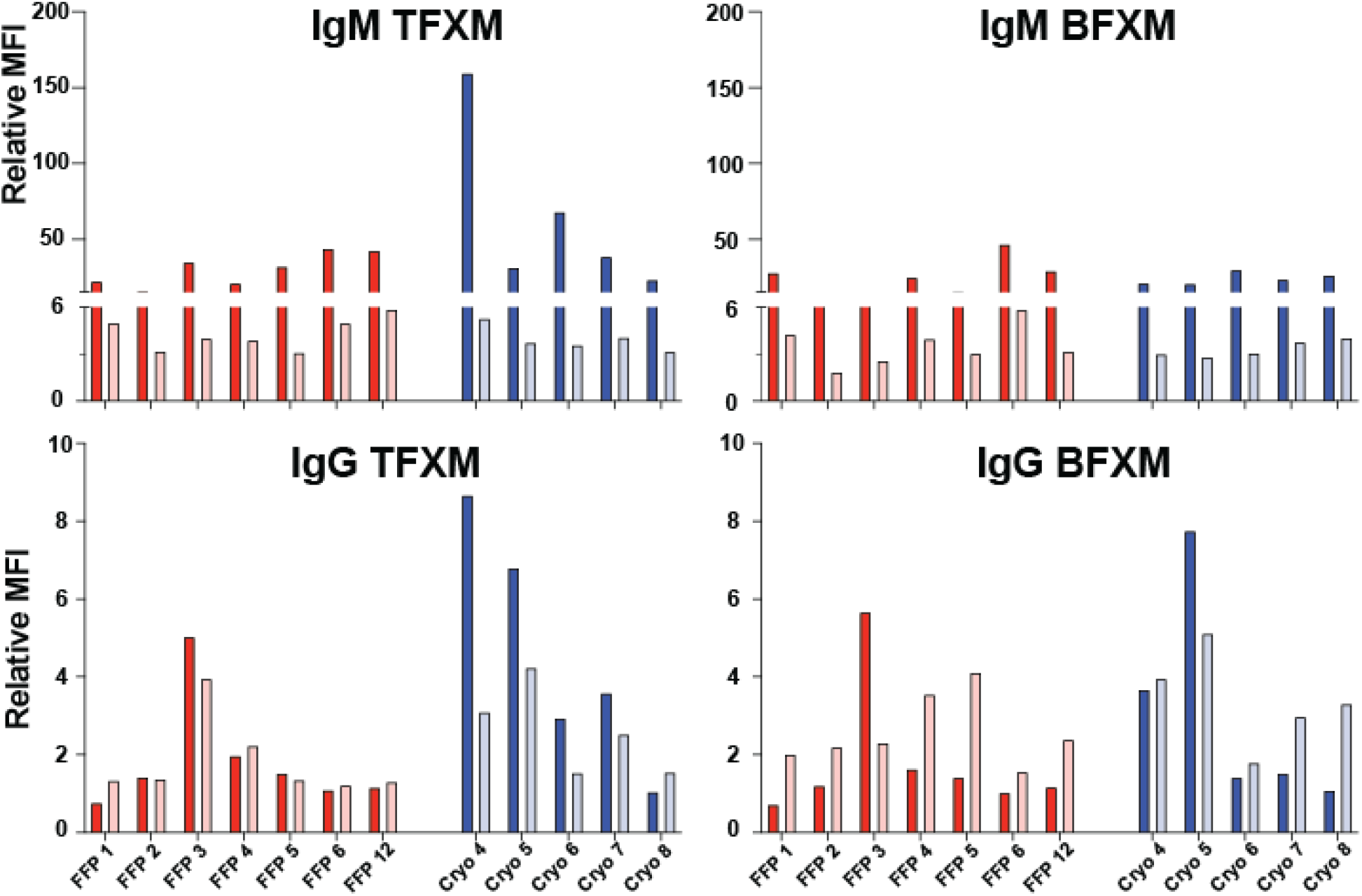
The IgM and IgG xenoantibody binding to CD3+ and CD21+ GTKO PBMCs of each individual blood product (FFP in red, cryo in blue). The dark bars represent neat sera samples and the light bars represent a 1:50 dilution. All values are depicted as MFI relative to negative control.

To evaluate the pathogenicity of the xenoantibodies in the blood products, a flow cytometry-based CDC assay was performed with or without DTT treatment **(Figure 3)**. Similar to the FCXM results, the degree of target cell cytotoxicity varied between specimens (mean cytotoxicity 40.30 ± 24.40%, p = 0.0484) with 6 samples having a CDC > 40% (3 FFP, 3 cryo). The addition of DTT to assess the effect of IgG resulted in a decrease in cell death (mean cytotoxicity 28.63 ± 20.69%, p = 0.3648) with only 4 samples (1 FFP, 3 cryo) having a CDC > 40% following DTT-treatment. There was no statistical difference between cytotoxicity with the addition of DTT (p = 0.2415).

**Figure 3:**
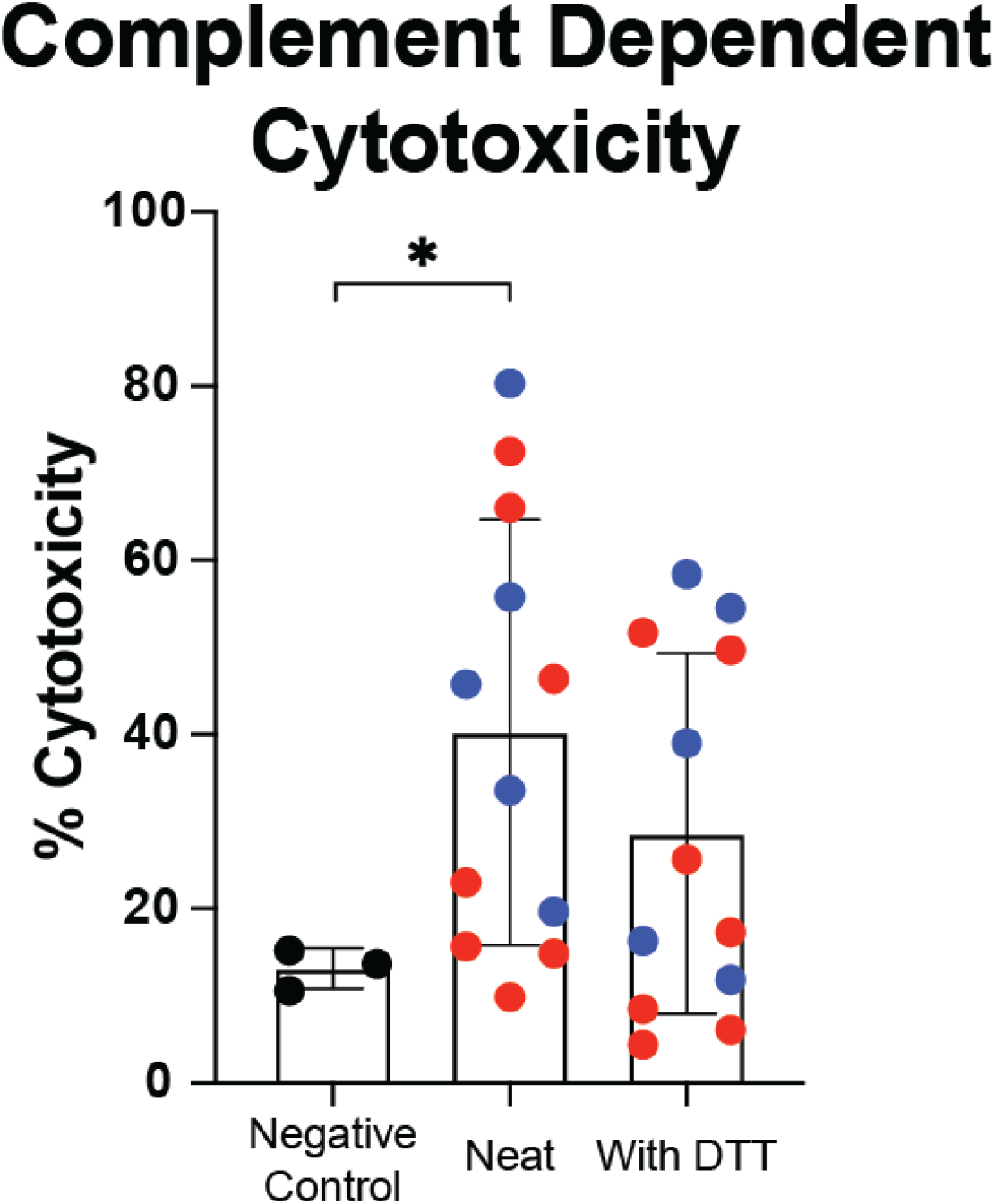
Blood product xenoantibodies are capable of complement-dependent cytotoxicity. The results of a flow cytometry-based CDC assay with neat (middle bar graph) and DTT-treated (right bar graph) FFP (red circles) and cryo (blue circles). Blood product xenoantibodies resulted in a significant amount of cellular cytotoxicity over negative control when neat sera was used (^*^, p = 0.0484) but not following DTT treatment (p = 0.3648). There was no significant difference between sera with and without DTT treatment (p = 0.2415).

## Discussion

We sought to investigate the presence of non-αGal xenoantibodies in blood products administered to treat coagulopathy in surgical patients – specifically fresh frozen plasma (FFP) and cryoprecipitate (cryo). We demonstrate that nearly all tested blood products exhibit some level of non-αGal IgM and IgG xenoantibodies capable of complement-dependent cytotoxicity, but the amount is variable by individual unit. Substantially higher levels of IgM xenoantibody binding were observed compared to IgG; however different secondary antibodies prevent a direct quantitative comparison. Nevertheless, levels of IgG xenoantibodies were sufficient to elicit cytotoxicity in the absence of IgM following DTT treatment. Interestingly, we did observe an increase in IgG binding following a 1:50 dilution and this may reflect a prozone effect or more likely binding interference caused by the high concentration of IgM xenoantibodies. Nevertheless, these findings suggest that prior to the administration to xenotransplant recipients, centers should consider screening blood products for potential donor pig xenoantibodies to prevent passive transfusion of harmful xenoantibodies. This is particularly relevant for studies involving decedent or clinical xenotransplantation cases using a GTKO-only GTKO background.

Our findings are consistent with the literature regarding xenoantibodies in IVIg by Yamamoto et al (8). The authors similarly reported a high level of non-αGal xenoantibody to GTKO red blood cells (RBCs) in five separate preparations of IVIg, but their findings report a dilutional effect in IgG xenoantibody binding at 1:20 that we failed to see at 1:50 (both publications saw a level of IVIg and blood product IgM xenoantibody binding that rapidly declined with dilution). Distinct from our report, the authors of the Yamamoto et al publication possessed triple knockout (TKO) pig cells. The authors were able to test their IVIg preparations on the TKO cells with reportedly no detectable xenoantibody binding, though this finding has since been debated (9). Nevertheless, the presence of xenoantibodies in blood products derived from human donors is unsurprising, as nearly all individuals possess some level of detectable non-αGal xenoantibodies. However, to date, no peer-reviewed report has documented their presence (22).

This study signifies a gradual shift in xenotransplantation: with multiple groups now demonstrating the ability to avoid hyperacute rejection, the focus is increasingly shifting to the perioperative management of xenograft recipients. This goes beyond the current focus on optimizing immunosuppression, expanding to encompass the proper perioperative management of post-surgical complications, such as bleeding. Furthermore, without appropriate testing or rationale, the assumption that the products intended for human allograft recipients are equally safe for xenograft recipients should be questioned. There are several important limitations to this study to address. First, blood products are a limited resource, and we were unable to obtain sufficient quantities of other blood products (e.g. platelets and packed red blood cells) for testing, though we plan to proceed with these studies in the future. Secondly, we were limited in the availability of TKO background cells for testing. Given the non-αGal xenoantibody target(s) detected in the blood products is unknown, and may include the Sda antigen, a TKO background may decrease blood product xenoreactivity. Nevertheless, some groups are choosing to pursue a GTKO background for their decedent and living recipients and these results carry relevance to their studies and patient care (23–26). Further studies are also required to determine the *in vivo* effect of these antibodies and whether their presence contributes to xenograft dysfunction or injury.

Xenotransplantation is approaching a pivotal and exciting phase, with promising results from early clinical implementations, a high demand for donor organs, and increasing enthusiasm for clinical trials. Despite these factors, the progression to a clinical trials should proceed cautiously, and assumptions regarding perioperative care must be thoroughly tested. One such assumption is that products or medications that historically proven safe for allografts recipients will likewise be safe for xenograft recipients (i.e. blood products). While this report demonstrates the existence of cytotoxic non-αGal xenoantibodies in both FFP and cryo, further work is needed to investigate the potential *in vivo* effect to a xenograft.

## Data Availability

All data produced in the present study are available upon reasonable request to the authors

## Abbreviation

αGal: galactose-α-1,3-galactose
AMR: Antibody-Mediated Rejection
BFXM: B cell flow crossmatch
DSA: Donor-Specific Antibody
FBS: Fetal Bovine Serum
FXM: Flow Crossmatch
GE: Genetically Engineered
GGTA1: Galactose-α-1,3-galactosyltransferase
HLA: Human Leukocyte Antigen
MHC: Major Histocompatibility Complex
MFI: Median Fluorescence Intensity
MHC: Major Histocompatibility Complex Neu5Gc, N-Glycolylneuraminic acid
PBMC: Peripheral Blood Mononuclear Cell
PBS: Phosphate Buffered Saline
SAB: single antigen beads
SLA: Swine Leukocyte Antigen
Tg: transgenic
TFXM: T cell flow crossmatch
TKO: Triple Knock Out

## Acknowledgement

The authors thank Kristin Whitworth (NSRRC) for coordinating GGTA/CD55Tg pigs for our study and the Duke University Blood Bank for their coordination and help with sample acquisition. The authors greatly appreciate the Duke Department of Laboratory Animal Research (DLAR) veterinarians for their support of the study with excellent animal care; and the Duke Surgery Training and Animal Research Core (STAR-C) staffs for their daily animal care; and the Duke University Clinical Transplantation Immunology Laboratory (CTIL).

## Author contribution

JML: Contributed to idea and hypothesis development, experimental design, and writing of the manuscript.

MH: Conducted in vitro experiments and participated in writing the manuscript.

JY, ZC: Assisted with in vitro experiments, sample acquisition, and critical review of the manuscript.

AJ, SK: Contributed to idea and hypothesis refinement, sample acquisition, and critical review of

JK: Contributed to idea and hypothesis refinement, sample acquisition, manuscript validation, and securing funding

## Data and Material Availability

The primary data that support the findings of this study are available from the corresponding authors upon reasonable request.

## REFERENCES

1. Cooper DKC. A brief history of cross-species organ transplantation. Proceedings (Baylor University. Medical Center) 2012; 25: 49.

2. Adams AB, Kim SC, Martens GR et al. Xenoantigen Deletion and Chemical Immunosuppression Can Prolong Renal Xenograft Survival. Annals of Surgery 2018; 268: 564.

3. Cooper DKC, Hara H, Iwase H et al. Justification of specific genetic modifications in pigs for clinical organ xenotransplantation. Xenotransplantation 2019; 26: e12516.

4. 2023 News - UM Medicine Faculty-Scientists and Clinicians Perform Second Historic Transplant of Pig Heart into Patient with End-Stage Cardiovascular Disease | University of Maryland School of Medicine [Internet]. [cited 2024 Sep 2] Available from: https://www.medschool.umaryland.edu/news/2023/um-medicine-faculty-scientists-and-clinicians-perform-second-historic-transplant-of-pig-heart-into-patient-with-end-stage-cardiovascular-disease.html

5. World’s First Genetically-Edited Pig Kidney Transplant into Living Recipient Performed at Massachusetts General Hospital [Internet]. Massachusetts General Hospital [cited 2024 Apr 1] Available from: https://www.massgeneral.org/news/press-release/worlds-first-genetically-edited-pig-kidney-transplant-into-living-recipient

6. First-Ever Combined Heart Pump & Gene-Edited Pig Kidney Transplant Gives New Hope to Patient with Terminal Illness | NYU Langone News [Internet]. [cited 2024 Sep 2] Available from: https://nyulangone.org/news/first-ever-combined-heart-pump-gene-edited-pig-kidney-transplant-gives-new-hope-patient-terminal-illness

7. Cooper DKC, Yamamoto T, Hara H, Pierson III RN. The first clinical pig heart transplant: Was IVIg or pig cytomegalovirus detrimental to the outcome? Xenotransplantation 2022; 29: e12771.

8. Yamamoto T, Cui Y, Patel D et al. Effect of intravenous immunoglobulin (IVIg) on primate complement-dependent cytotoxicity of genetically engineered pig cells: relevance to clinical xenotransplantation. Scientific Reports 2020; 10: 11747.

9. Mangiola M, Stern J, Kim J, et al. Anti-pig antibodies in human immunoglobulin preparations. IPITA-IXA-CTRMS 2023 - Virtual [Internet]. IPITA-IXA-CTRMS Joint Congress 2023 [cited 2024 Sep 2] Available from: https://app.sandiego2023.org/virtual/lecture/422

10. Kotter JR, Drakos SG, Horne BD et al. Effect of Blood Product Transfusion– Induced Tolerance on Incidence of Cardiac Allograft Rejection. Transplantation Proceedings 2010; 42: 2687.

11. Early risks of a heart transplant [Internet]. Organ transplantation - NHS Blood and Transplant [cited 2024 Sep 2] Available from: https://www.nhsbt.nhs.uk/organ-transplantation/heart/benefits-and-risks-of-a-heart-transplant/risks-of-a-heart-transplant/early-risks-of-a-heart-transplant/

12. Sniecinski RM, Levy JH. Bleeding and management of coagulopathy. The Journal of Thoracic and Cardiovascular Surgery 2011; 142: 662.

13. Stanworth SJ. The Evidence-Based Use of FFP and Cryoprecipitate for Abnormalities of Coagulation Tests and Clinical Coagulopathy. Hematology 2007; 2007: 179.

14. Li M, Cardigan R, Thomas S, Vassallo R. Production and Storage of Blood Components [Internet]. In: Practical Transfusion Medicine. John Wiley & Sons, Ltd, 2022 [cited 2024 Sep 2] : 278.Available from: https://onlinelibrary.wiley.com/doi/abs/10.1002/9781119665885.ch24

15. Blumberg N, Gettings KF, Heal JM. ABO Mismatched Platelet, Plasma and Cryoprecipitate Transfusions and Bleeding in Transfused Surgical Patients. Blood 2005; 106: 4166.

16. An association of ABO non-identical platelet and cryoprecipitate transfusions with altered red cell transfusion needs in surgical patients - Refaai - 2011 - Vox Sanguinis - Wiley Online Library [Internet]. [cited 2024 Sep 2] Available from: https://onlinelibrary.wiley.com/doi/10.1111/j.1423-0410.2010.01464.x

17. Martens GR, Reyes LM, Butler JR et al. Humoral reactivity of renal transplant-waitlisted patients to cells from GGTA1/CMAH/B4GalNT2, and SLA class I knockout pigs. Transplantation 2017; 101: e86.

18. Ho C-S, Lunney JK, Lee J-H et al. Molecular characterization of swine leucocyte antigen class II genes in outbred pig populations. Animal Genetics 2010; 41: 428.

19. Ho C-S, Lunney JK, Franzo-Romain MH et al. Molecular characterization of swine leucocyte antigen class I genes in outbred pig populations. Animal Genetics 2009; 40: 468.

21. Ladowski JM, Martens GR, Reyes LM et al. Examining the biosynthesis and xenoantigenicity of class II swine leukocyte antigen proteins. Journal of Immunology 2018; 200: 2957.

22. Ladowski JM, Chapman H, DeLaura I et al. Allosensitisation in NHP results in cross-reactive anti-SLA antibodies not detected by a lymphocyte-based flow cytometry crossmatch. HLA 2024; 104: e15599.

23. Kim J, Stern J, Khalil K, et al. Cryoprecipitate and Fresh Frozen Plasma Xenoantibody Testing. ATC 2024 Poster Abstracts. American Journal of Transplantation 2024; 24: S562.

24. Porrett PM, Orandi BJ, Kumar V et al. First clinical-grade porcine kidney xenotransplant using a human decedent model. American Journal of Transplantation 2022; 22: 1037.

25. Montgomery RA, Stern JM, Lonze BE et al. Results of Two Cases of Pig-to-Human Kidney Xenotransplantation. New England Journal of Medicine 2022; 386: 1889.

26. Loupy A, Goutaudier V, Giarraputo A et al. Immune response after pig-to-human kidney xenotransplantation: a multimodal phenotyping study. Lancet (London, England) 2023; 402: 1158.

27. Judd E, Kumar V, Porrett PM et al. Physiologic homeostasis after pig-to-human kidney xenotransplantation. Kidney International 2024; 105: 971.

